# Medication Management at Home and Medication Adherence in Older Patients with Polypharmacy: In-Depth Interviews with Home Visits

**DOI:** 10.1101/2021.09.15.21263068

**Authors:** Pasitpon Vatcharavongvan, Viwat Puttawanchai

## Abstract

**Background:** Older patients with multiple non-communicable diseases (NCDs) usually require ≥5 concurrent medications or polypharmacy. Medication adherence is the main concern in these patients. Medication management at home may play an important role in medication adherence; yet, unlike other factors, the topic has not been well examined.

**Objective:** This study aims to explore how medication management at home affects medication adherence qualitatively.

**Methods:** We conducted home visits and in-depth interviews with 19 patients aged ≥60 years with polypharmacy and took photos of medication storage locations. Transcripts were analyzed using thematic content analysis.

**Results:** Of the 19 patients (mean age=69 years), nine reported good medication adherence. Two themes emerged: medication management at home and factors affecting medication adherence. Medication management at home comprised three subthemes: a medication storage system, a medication sorting system, and remaining medications. Some patients with medication nonadherence removed medications from blister packages. Other factors affecting medication adherence included knowledge, attitude, and lifestyles. All the patients had a positive attitude towards medication adherence; however, misunderstanding about medication administration prevented them from adhering to medications.

**Conclusion:** Medication management at home for the elderly with polypharmacy affected medication adherence. Health professionals should explore how patients manage medications at home and their knowledge about medication administration to improve medication adherence.

**Key messages:**

- Older patients had their own medication management system.
- Removing medications from blister packages was associated with medication nonadherence.
- Unintentional medication nonadherence was common in older patients engaging in outdoor activities.

## Introduction

Thailand has been an aging society since 1997 (1). According to the Elderly Person Act, 2003, older adults are those aged 60 years or more. In Thailand, the proportion of older adults has increased, from 10.7% in 1997 to 14.9% in 2013 (1). Older Thai adults are estimated to reach 1.7 million (26.6%) in 2030 (2). Common health problems in this age group are noncommunicable diseases (NCDs), such as diabetes mellitus and hypertension, and their complications, such as ischemic heart disease and stroke. With the increasing number of older adults, the prevalence of NCDs is expected to rise. Data from 2014 indicate that approximately two million (18.3%) Thai older adults had one or more NCDs (3). According to standard guidelines for NCDs, such as diabetes mellitus (4)and hypertension (5), patients with NCDs are prone to take multiple medications. Those who develop complications may need more than ten medications to treat diseases and prevent further complications (6).

The concurrent use of multiple medications, or polypharmacy, among older adults is a global phenomenon that has been reported in several countries and various settings (7-10). For example, 20% of older Scottish adults used five or more medications, and 5.8% used more than ten medications (7). Polypharmacy in Thailand is not well studied; however, the findings report a high prevalence of polypharmacy (11, 12). Older Thai patients are at risk of polypharmacy because of NCDs and a national health policy that allows patients to access multiple medical doctors.

Older adults are at risk of poor medication adherence, and several factors determining medication adherence have been reported (13, 14). Yap and Kwan (13) conducted a systematic review examining barriers to medication adherence. A total of 80 factors were identified and grouped into five categories: patient, medication, physician, system-based, and other factors (13). However, this review does not mention the magnitude of these identified barriers to medication adherence. Another factor associated with medication adherence in older adults is medication management at home (14). Medication management at home includes, for example, medication storage locations, the presence of expired medication, the retention of discontinued medication, hoarding of medications, and the presence of duplicated medications. In their study, Sorensen et al. (14) found that having duplicated medications (p<0.0001), retaining expired medication (p=0.020), and storing medications in multiple locations (p=0.025) were related to poor medication adherence.

Our study qualitatively explored medication management at home among older patients with polypharmacy in a primary care setting. This study aims to help illuminate how these patients manage their medications at home and how medication management at home is related to medication adherence.

## Methods

### Design and setting

Based on interpretivism, we examined medication management at home in patients with polypharmacy between May and June 2015 in the catchment area of the Kukot Primary Care and Applied Thai Traditional Medicine Center (hereafter Kukot PCU), which is located in Kukot municipality, a semiurban area in Pathum-Thani province, in the central region of Thailand. Interpretivism allows researchers to understand the interpretation of participants’ worldviews (15). The Kukot PCU is a primary care unit with doctors, nurses, and pharmacists providing primary care services to the community. We conducted in-depth interviews, visited the patients’ houses, and took photographs of medication storage locations. All personal data were kept strictly confidential.

### Participants

We recruited patients from the Kukot PCU. These patients were eligible if they 1) were 60 years old or older; 2) were diagnosed with two or more NCDs, including type 2 diabetes mellitus, hyperlipidemia, hypertension, ischemic heart disease, chronic heart failure, cerebrovascular disease, and epilepsy; and 3) concurrently used five or more medications for NCDs. The patients were invited by telephone to participate in interviews and home visits with their preferred dates and time. The purposes of the interviews and home visits were provided, and all patients’ concerns regarding the study were addressed.

### Data collection

Data were collected from in-depth interviews and photos of medication storage locations. The first author (PV) conducted in-depth interviews at the patients’ houses. The interviews were digitally audio-recorded with permission from the patients. A semistructured questionnaire was developed to explore three topics: (a) the patients’ history of NCDs, (b) medication-related problems, and (c) medication management at home. Each interview lasted approximately 60 to 90 minutes. After the interviews, PV asked the patients to show their medication storage locations, and with their permission, took photos of medications in their storage locations. The patients’ reasons for storing medications in each location were explored. We took notes during the interviews and drew sketches of the medication storage locations. At the end of each interview, PV summarized the information and asked each participant to correct any mistakes or misunderstandings.

### Analysis

Content analysis was conducted without software. Content analysis is a systematic analytical method (16). The analytical process began soon after the first few interviews. The interviews were transcribed. The authors reviewed all audio records and transcripts. Photos and notes were used to complement the data analysis. The photos illustrated the storage locations and how medications were stored at each location, which helped us recall our observations and expanded our understanding of each participant’s medication storage system. Themes and subthemes related to medication management at home were generated. The authors independently applied codes to all themes and subthemes. We then iteratively organized and categorized the themes and subthemes. The authors discussed resolving the dissimilarity of codes, themes, and subthemes. A diagram was drawn to explain how the patients managed their medications at home.

We compared medication management at home between those with good adherence and those without adherence. The patients with medication nonadherence were those who (a) took medications at the wrong time, (b) missed medications, or (c) had intentionally stopped medications at least once within the two weeks before the interviews. We defined the patients with good control of their NCDs as those with one of the following: (a) fasting blood sugar between 90-130 mg/dL during the last two visits, (b) systolic and diastolic blood pressure lower than 140 and 90 mmHg, respectively, during the last two visits, and (c) no hospital admissions due to NCDs within the previous six months.

## Results

Of 23 invited patients, four did not want to participate. Of the 19 patients, 11 were women, and 9 reported good medication adherence (Table 1). Their median age was 69 years. Diabetes mellitus and hypertension were the two most common diagnoses. The patients used six medications on average, stored medications in one to four locations, and regularly visited one to four doctors with different specialties. Most of the patients managed their medications themselves. Only two received help from their relatives because one had visual impairment and another had dementia with impaired cognitive function.

**Table 1.** The patients’ characteristics and medication management at home

| Patients | Age | Gender | Diseases | Medications (n) | Locations (n) | Medication management | RfP/RfB | Doctor(s) |
| --- | --- | --- | --- | --- | --- | --- | --- | --- |
| Good adherence |  |  |  |  |  |  |  |  |
| 1 | 70-74 | Male | HT, DLP | 5 | 2 | Self | RfB | 1 |
| 2 | 80-84 | Male | HT, DLP | 5 | 1 | Self | No | 1 |
| 3 | 60-64 | Male | DM, HT | 6 | 3 | With relative | RfBP | 1 |
| 4 | 60-64 | Male | DM, HT | 5 | 3 | Self | RfB | 1 |
| 5 | 70-74 | Female | DM, HT | 6 | 3 | Self | No | 1 |
| 6 | 60-64 | Female | DM, HT, CHF | 10 | 3 | Self | No | 4 |
| 7 | 70-74 | Female | DM, HT | 7 | 3 | Self | RfB | 1 |
| 8 | 60-64 | Female | DM, HT | 9 | 2 | Self | No | Unknown |
| 9 | 70-74 | Female | DM, HT, DLP | 7 | 3 | Self | No | 1 |
| Nonadherence |  |  |  |  |  |  |  |  |
| 1 | 65-69 | Female | DM, HT | 5 | 3 | Self | RfB | 1 |
| 2 | 70-74 | Male | DM, HT | 8 | 3 | Relative | No | 4 |
| 3 | 60-64 | Male | DM, DLP | 5 | 4 | Self | RfBP | 1 |
| 4 | 85-89 | Female | HT, DLP | 5 | 2 | Self | RfBP | 1 |
| 5 | 70-74 | Male | DM, HT | 6 | 2 | Self | No | Unknown |
| 6 | 60-64 | Female | DM, HT | 5 | 2 | Self | RfBP | 2 |
| 7 | 70-74 | Male | DM, HT | 8 | 3 | Self | No | 1 |
| 8 | 70-74 | Female | DM, HT, DLP | 5 | 2 | Relative | No | 1 |
| 9 | 70-74 | Female | DM, HT | 5 | 3 | Self | RfBP | 1 |
| 10 | 65-69 | Female | DM, HT, DLP | 6 | 2 | Self | No | Unknown |

Abbreviations: RfBP: removed from blister packs, RfB: removed from bags, DM: diabetes mellitus, HT: hypertension, DLP: dyslipidemia, CHF: congestive heart failure

Patients explained how they stored and organized medications at home, how they managed remained medications, and how their knowledge, attitude, and lifestyles affected medication adherence (Table 2).

**Table 2.** Coding tree: Themes, subthemes, and key findings from the interviews

| Themes and subthemes | Key findings |
| --- | --- |
| Medication management at home |  |
| Medication storage system | Stored newly received medications in one location<br>Stored medications for use in one to two locations<br>Stored insulin in the refrigerator |
| Medication sorting system | Most handled medications by themselves<br>Some used remaining medications before newly received medications<br>Medications were handled in three patterns: (1) kept in the same bags, (2) removed from the bags, and (3) removed from the blister packs<br>The medication handling system was not associated with medication adherence |
| Remaining medications | The poorly-controlled group had more remaining medications than the other group<br>The remaining medications were (1) kept, (2) shared, (3) thrown away, or (4) returned to Kukot PCU |
| Other factors affecting medication adherence |  |
| Knowledge and attitudes about medications and diseases | Knowledge and attitude affected medication adherence<br>Misunderstandings about the timing and benefits of medications were related to medication adherence |
| Lifestyle | Those who still worked or were socially active were at risk of poor medication adherence |

### Medication management at home

#### Medication storage system

The patients stored medications in one to four locations. Generally, at least two locations were used: one for newly received medications and one based on where the medications were taken (Figure 1). Only one patient kept all medications in one pouch. The newly received medications were kept in the bags provided by the Kukot PCU and stored in different locations, such as under a bed, on a wardrobe, and on a hook. Some patients had more than one location based on where medications were taken. For example, one patient kept glipizide in a kitchen to be taken before a meal in the morning and kept other after-meal medications on a dining table. A refrigerator served as an additional storage location for insulin. The number of storage locations was similar between those with good medication adherence and those with medication nonadherence.

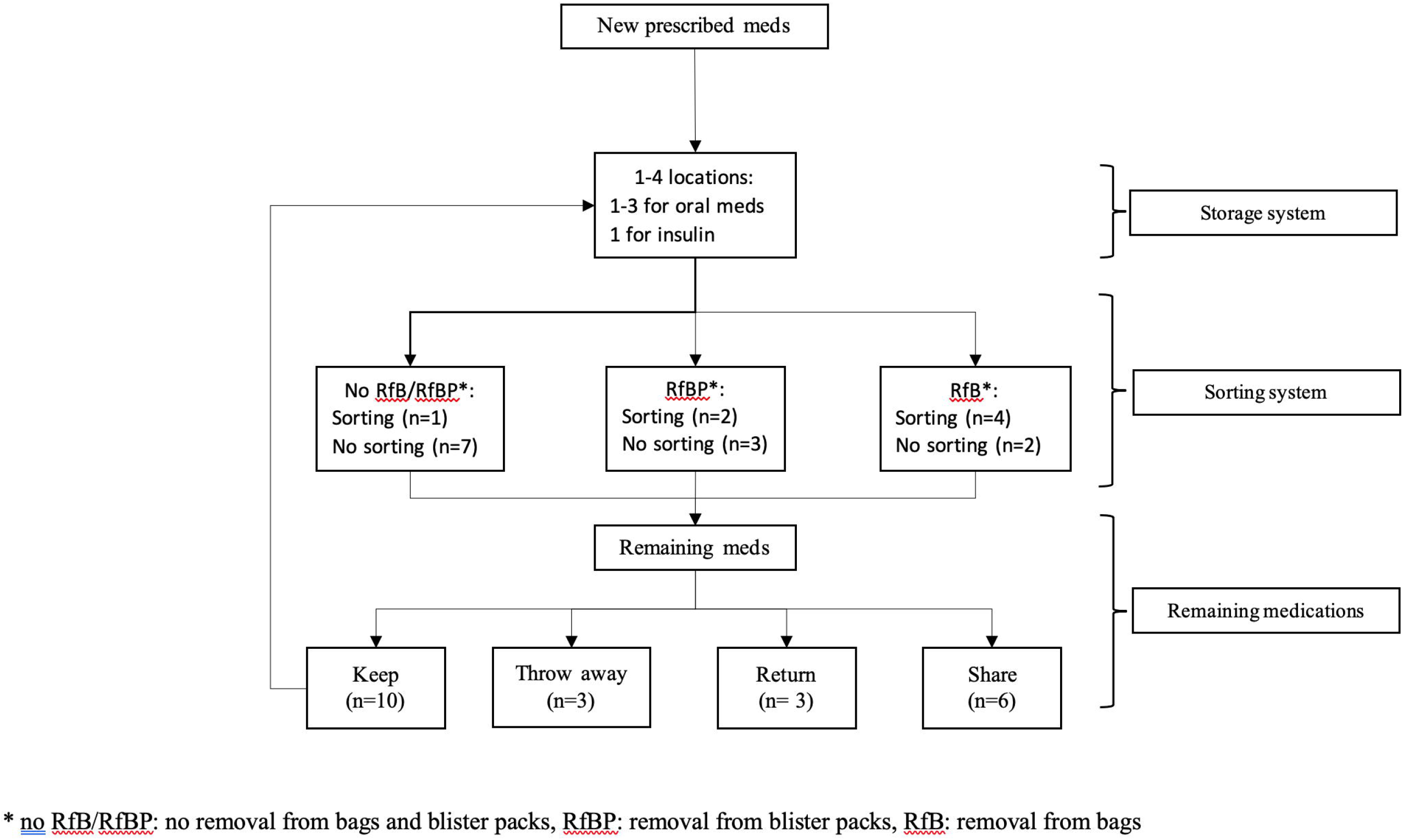

> “[I kept medications] in a drawer. [Some] are hung…on a bed…because I believe the room is not hot and moist. Some medications should not be kept in a hot and moist room, should they? I organize them all myself. A doctor told me to keep them in a dry and cool location.” (male)
>
> “Insulin is kept in a fridge. These medications are in a container, this container… they are for both diabetes and hypertension.” (male)

Medications were placed in one to two locations depending on the patients’ lifestyles. These locations included the kitchen, dining room, bed, living room, and work area. These medications were kept in plastic bags, plastic containers, and baskets. Only one patient kept medications in a medication storage cabinet. Almost half of the patients in both the good adherence and nonadherence groups used remaining medications before starting newly received medications. Few patients knew their medications’ expiration dates.

> “I use remaining medications first. I do not look at an expiratory date… [I look at] the date I received the medications and use the older ones first.” (female)
>
> “I keep medications in the bedroom… and hang some on a bookshelf. Medications taken in the morning are kept in the kitchen… both before and after a meal. There is a dining table there in the kitchen.” (male)

#### Medication sorting system

Most patients organized medications themselves because they had good vision and cognitive function or did not want to disturb their relatives. Only two patients had their relatives help manage their medications because of visual and cognitive impairment.

> “I remember all medications because I take them every day. I know which medications have to be taken before a meal in the morning, and I take them before breakfast.” (male)
>
> “My vision and hearing are not that good… I can’t read labels. My relative writes instructions [for how to take them] in big letters… Sometimes, she reads the labels. She tells me to take these medications after breakfast… I need her help… She doesn’t organize them every day. Medications from PCU are put in baskets… one for morning and the other for the evening.” (male)
>
> “I don’t want my relative to help me because I can do it myself. I don’t want to disturb him.” (female)

In everyday practice at the Kukot PCU, patients receive medications in blister packs from pharmacists. All blister packs of the same medications were placed in the same plastic bags with prescription labels. Little is known of what they do with medications when they are at home. From the interviews, we found three patterns the patients used to sort medications in both the good adherence and nonadherence groups (Figure 1).

The first pattern was to put small bags of the same blister packs together (no RfB/RfBP pattern). In this pattern, medications remained in their blister packs, and the packs remained in the small bags received from health care providers. The second pattern was to take some blister packs out of the small plastic bags and put the packs of different medications together in other containers (RfB pattern). The last pattern was to remove medications from blister packs and put them in containers with or without sorting (RfBP pattern). There was no specific reason given for the pattern the patients used. They just said, “It is convenient for me.” The patients sometimes changed from one pattern to another. For example, some patients started with the second pattern and then shifted to the last pattern, so they did not have to remove medications from blister packs every time they took them. Medication nonadherence was more common among those with RfBP than among those with the other two patterns.

> “I cut packages and put them back in small bags. The medications are still in the packages. I mostly do it at night, so I can take the medications tomorrow.” (male)
>
> “I have a box. I put one pack at a time in the box in the evening. When the medications are almost gone, I put another one in there.” (female)

#### Remaining medication management

At each follow-up visit, some of the patients had leftover medications, which were managed in four ways: keep, share, return, or throw away (Figure 1). Most patients kept their leftover medications and used them either before or after starting newly received medications. The main reason for keeping leftover medications was that the patients did not want to waste them. Few patients knew their medications’ expiration dates.

> “I don’t know where I can find an expiratory date, but I normally finish the remaining medications before taking newly received medications. I don’t want to waste the medications.” (female)

Less than half of the patients shared leftover medications, returned them to the Kukot PCU, or threw them away. Those who shared their leftover medications shared them with relatives and neighbors. The patients explained that giving medications to others was “better than throwing them in the trash” and “like a donation.” One patient experienced adverse consequences of medication sharing happening to a recipient and never shared medications again. Some patients used more than one method to manage leftover medications.

> “My neighbor has diabetes. Once his medications ran out. He asked me for medications. I had plenty of them, so I gave him ten packages… approximately 100 pills. He was delighted and thanked me for the medication.” (female)
>
> “I used to share metformin with my relative, and he got severe diarrhea. After that, I never shared medications with anyone again.” (female)

### Factors affecting medication adherence

Factors affecting medication adherence emerged during the interviews and complemented the findings of medication management at home. These factors included 1) knowledge and attitudes toward diseases and medications and 2) patients’ lifestyles.

#### Knowledge and attitudes

Knowledge about medications was associated with medication adherence. Misunderstanding the timing and benefits of medications resulted in poor medication adherence. For example, some patients believed that medications labeled “take with food” had to be taken “after a full meal,” although some medications, such as antihypertensive agents, could be taken without a meal. In Thai culture, a meal is a plate or bowl filled with rice or noodles. Even if the patients had a cup of coffee and biscuits, they chose to wait until they finished a “full meal” to take medications. Some patients stated that “[medications to be taken] after dinner can be taken before bedtime.” A few patients took medications “depending on symptoms.” For example, one patient injected insulin in the evening only when he had polyuria.

Attitudes regarding the benefits and harms of medications were similar between the good adherence and nonadherence groups. All the patients believed that medications helped control blood pressure, cholesterol levels, and blood sugar levels. Some were afraid that without medications, they would develop complications. The patients were aware of the side effects of medications and overcame their fear by recognizing the benefits of the medications.

> “If I am afraid of side effects or if I don’t take medicines, I may have complications from diseases… I have to accept that fact. Take them. If there will be side effects, let them be. I am not worried at all.” (male)

Most patients believed they had to take medications for the rest of their lives. Their explanations included diseases that were not curable, fear of having complications, and the experiences of their relatives who developed complications. Some patients wanted to take fewer medications than they were currently taking. All the patients trusted their doctors’ decisions regarding prescribed medications. They would not discontinue medications unless their doctors directed them to do so.

> “I can’t stop taking these medications. Sugar and blood pressure go up and down all the time. Without medications, I can’t control them.” (female)

Knowledge or misunderstandings about medication administration seemed to have a greater effect on medication adherence than attitudes toward medications. The patients showed their willingness to adhere to prescriptions; however, their misunderstandings about medication administration resulted in medication nonadherence.

#### Lifestyle: Work and outings may affect medication adherence

The patients’ lifestyles were related to medication adherence. Most patients spent most of their time at home, while some still worked or regularly visited their relatives. For the latter group, medication nonadherence was found among those who did not prepare their medications in advance. A few patients who still worked forgot to take medications after lunch.

> “I rarely miss medications, especially when I stay at home. The exception is when I go out; I sometimes forget to take medications with me.” (male)

## Discussion

This study aimed to explore how patients manage medications at home, which can be complicated for those with polypharmacy. The way patients manage medications at home may affect medication adherence. We interviewed the patients at their houses and photographed their medication storage locations. The patients were those aged 60 years or older with polypharmacy who had visited Kukot PCU in the past year. In general, two themes emerged: medication management at home and factors affecting medication adherence. The first comprised three subthemes: 1) medication storage system, 2) medication sorting system, and 3) remaining medication management. The latter comprised two subthemes: 1) knowledge and attitudes and 2) lifestyle.

Few studies have explored medication storage systems, particularly in those with polypharmacy. The medication storage system is the way the patients store their medications at home. A previous study found that patients with multiple NCDs and polypharmacy usually stored medications in two locations (11). Our findings illuminated the purposes behind storage locations for newly prescribed medications and those based on where medications were taken. Medication storage locations for the latter purpose depended on the patients’ lifestyle and knowledge about appropriate medication storage. While other studies indicate that multiple storage locations are associated with medication nonadherence (14) and poor health outcomes (17), our study did not find a difference in storage locations between patients with good adherence and those with medication nonadherence. Although storage locations seemed not to be associated with medication adherence, asking patients about how they spend their time at home instead of their number of storage locations may help doctors give patients appropriate advice regarding how to store medications properly.

Patients might take newly received or leftover medications first. Either way, our study found that few patients knew their medications’ expiration dates, which put them at risk of having expired medications. Sorensen et al. (17) found that patients with polypharmacy had a high chance of having expired medications. Another study further illustrated that having expired medications at home was associated with retaining discontinued medication, medication nonadherence, and having multiple storage locations (1). Educating patients about expiration dates or making them visible, such as pictorial aids (18), will prevent older patients from ingesting expired medications.

Most of the patients managed and sorted their medications themselves (11). Our study found that the patients’ vision, cognitive function, and consideration of their relatives played an important role in their decision to seek help from relatives with sorting medications. The findings suggest that asking patients directly about medication management at home will give doctors sufficient and accurate information. Most nonadherence was associated with the removal of medications from blister packs. This finding was congruent with a previous study that reported the association between a lack of medication box use and medication nonadherence (13). We urge that patient education programs should include an effective medication sorting system to improve medication adherence among older patients, as it is not emphasized in most practice guidelines for diabetes mellitus (19), dyslipidemia (20), and hypertension (21).

The universal health coverage scheme covers most patients at the PCU; hence, they do not need to pay for refilled medications. A previous study found that 37% of those with polypharmacy retained discontinued medications, and 10.5% had expired medications (11). The findings reflect the fact that patients had remaining medications at the time they received medication refills. Our study confirmed this assumption and further illustrated that most patients kept remaining medications to use either before or after newly received medications. As previously mentioned, the most concerning finding was the patients’ unawareness of medication expiration dates. A previous study revealed an association between having expired medications and medication nonadherence (11). It is very important to remind and educate patients to look at expiration dates before taking the remaining medications. The less common ways of managing leftover medications included medication sharing, returning medications, and disposing of medications. Approximately 15% of older patients with polypharmacy share medications with other people (11).

Other factors that affect medication adherence were knowledge and lifestyles. Their misunderstandings of medication administration derailed their intentions to adhere to medications. For example, antihypertensive agents were labeled with instructions to take them after a meal, which the patients understood as a meal with rice or noodles. They skipped the medication when they had not eaten a meal, such as breakfast, that day. The findings suggest that doctors should explore patients’ knowledge about medication administration and correct misunderstandings to improve medication adherence.

Unintentional medication nonadherence referred to medication nonadherence due to the inability to access medications, forgetting, interruptions of routine, and a lack of reminders (22). In our study, forgetting, interruptions of routines, and a lack of reminders among the patients, particularly those with good adherence, resulted in unintentional nonadherence. Using a pillbox and promoting family engagement in taking medications may improve patient medication adherence (23, 24).

### Strengths and limitations

There are some limitations to this study. First, medication adherence was based on interviews, which had a risk of false positives and lack of sensitivity because of recall biases and distortion of reality (25). The latter is a situation where a patient falsely reports good medication adherence to a doctor. Second, most patients managed their medications themselves because they had good vision and cognitive function; therefore, the findings cannot be applied to patients who receive help from other people. Last, medication management at home may change over time. One-time interviews could not capture such changes, which may have led to misinterpretation. Our findings, therefore, must be interpreted cautiously.

## Conclusions

Medication management at home in older patients with polypharmacy affected medication adherence. The findings encourage the use of a pillbox and discourage the removal of medications from bags and blister packages without using a pillbox. A patient’s misunderstanding about medication management should be explored and corrected.

## Data Availability

Data available upon request.

## Declarations

### Ethics approval and consent to participate

This study obtained ethical approval from the ethical review board for human research, Faculty of Medicine, Thammasat University (MTU-EC-CF-2-072/57). Written informed consent was obtained from all individual patients included in the study before the interviews.

### Consent for publication

Not applicable

### Availability of data and material

The data that support the findings of this study are not openly available due to patients’ confidentiality and are available from the corresponding author upon reasonable request.

### Competing interests

The authors declare that they have no competing interests

### Funding

Thailand Research Fund [Grant No TRG5780025]

### Authors’ contributions

Pasitpon Vatcharavongvan designed the study, collected data, analyzed the data, wrote the manuscript, and submit the manuscript. Viwat Puttawanchai designed the study, analyzed the data, and wrote the manuscript.

## Acknowledgment

We are grateful to all the patients who devoted their time to the interviews and their warm welcome during the interviews. We also thank the nurse practitioners at the PCU for contacting the patients and navigating our way to their houses. We are grateful to the director of Kukot PCU, who allowed us to conduct research in the catchment area. We thank the Thailand Research Fund [Grant No TRG5780025] for financially supporting our research project.

## Notes

### Competing Interest Statement

The authors have declared no competing interest.

### Author Declarations

This study obtained ethical approval from the ethical review board for human research, Faculty of Medicine, Thammasat University (MTU-EC-CF-2-072/57).

